# Blood Pressure and Heart Rate Stationarity in Predicting TEVAR Response

**DOI:** 10.64898/2026.10.12.26365335

**Authors:** Amira Alakhdar, Newell R. Washburn, Luka Pocivavsek, Nicole L. Bohr

**Affiliations:** Department of Chemistry, Carnegie Mellon University, Pittsburgh, PA 15213, USA; Department of Biomedical Engineering, Carnegie Mellon University, Pittsburgh, PA 15213, USA; Department of Surgery, University of Chicago, Chicago, IL 60637, USA; Department of Nursing Research and Evidence-Based Practice, UChicago Medicine, Chicago, IL 60637, USA

## Abstract

**Background:** Hypertension is a major risk factor for type B aortic dissections and rupture, with guidelines recommending systolic blood pressure (SBP) control below 130 mmHg to improve outcomes. While post-TEVAR blood pressure is often managed with a focus on mean SBP, little is known about temporal hemodynamic patterns and their association with surgical success. This study investigates blood pressure and heart rate (HR) time-series metrics within 24 hours post-TEVAR and their predictive potential for outcomes.

**Methods:** Blood pressure (n = 560) and HR (n = 680) time-series data from 31 patients were analyzed alongside preoperative aortic geometry metrics. Exploratory data analysis identified correlations between BP variability and outcomes. Stationarity tests were performed on time-series data to assess statistical properties. Logistic regression models incorporating hemodynamic metrics, along with CT-derived features, were used to predict TEVAR outcomes.

**Results:** Successful outcomes correlated with greater diastolic blood pressure variability and strictly stationary SBP. Logistic regression models combining these metrics along with heart rate standard deviation and preoperative aortic geometry achieved 96.67% 5-fold cross-validation accuracy in predicting outcomes.

**Conclusion:** Post-TEVAR behaviors of hemodynamic variables are critical predictors of TEVAR success, with higher DBP standard deviation and strict stationarity of SBP significantly associated with favorable outcomes. These findings highlight the importance of hemodynamic variability and stationarity in optimizing post-TEVAR patient care.

## Introduction

Aortic dissection (AD) is the most common fatal event involving the aorta and occurs in 5 to 30 cases per million of the population, resulting in approximately 12,000 deaths annually in the United States.^1^ In an acute AD, blood penetrates the aortic wall at the site of an intimal tear and proceeds to separate the aortic layers and extend up or down the aorta, creating a false lumen. This creates a problem of mechanical failure as the aorta, a composite pressurized tube, undergoes interfacial fracture. While surgical management has rapidly evolved in the advent of endovascular therapies, understanding this problem requires knowledge of mechanical factors such as aortic wall stress, aortic shape, and aberrant blood flow patterns.^2–5^ As a result, AD is a mechanical problem best understood through the use of geometry and hemodynamics.

Hypertension is a well-known risk factor for the propagation of type B aortic dissections and rupture. Clinical practice guidelines suggest maintaining a systolic blood pressure (SBP) below 130 mmHg for all patients to improve aortic outcomes and overall mortality.^6^ This is typically monitored in the outpatient setting using a single blood pressure reading that is known to vary substantially from visit to visit due to random variation around the patient’s “true” blood pressure owing to measurement errors.^7^ There is limited data on blood pressure immediately following surgical and/or endovascular repair, resulting in attempts to meet the guideline-directed blood pressure target. However, the more frequent collection of data could provide more information leading to a better understating of the behavior of blood pressure following endovascular repair, especially in the most critical 24 hours post-surgery. While temporal patterns are complex^8^, metrics such as blood pressure variability have been associated with higher risk of cardiovascular events and mortality. In fact, increased visit-to-visit blood pressure variability is associated with worse cardiovascular disease outcomes independent of mean blood pressure and other cardiovascular disease risk factors.^9, 10^ Such metrics have not been explored in AD where postoperative management has largely focused on reducing SBP.

This also holds true in examining heart rate. While there is more focus on blood pressure management in the long-term management of AD, clinical practice guidelines recommend use of beta blockers in the immediate phase to maintain a heart rate less than 60 in an effort to decrease aortic wall stress and prevent dissection-related events.^6^ An elevated resting heart rate has long been observed as a predictor of cardiovascular disease and cardiovascular mortality^11^; however, the concern surrounding potential for measurement error has led to more extensive examination of heart rate variability. As seen with blood pressure, increased visit-to-visit heart rate variability is independently associated with a higher risk of cardiovascular events.^12^ Furthermore, a recent study established that heart rate variability is positively associated with carotid intima-media thickness, which is a sign of early arteriopathy.^13^

Therefore, the purpose of this study is to analyze the behavior of blood pressure and heart rate (HR) time-series data in the 24-hour post-TEVAR by pairing unique blood pressure metrics with preoperative aortic geometry, which has been shown to predict post-TEVAR outcomes.^14^ This approach allows us to identify which hemodynamic metrics are most effective in predicting TEVAR outcomes.

## Methods

### Data Collection

A retrospective observational study was conducted using electronic medical record data at a single academic tertiary care facility; institutional review board approval was obtained for the conduct of this research. All patients who underwent TEVAR, had a minimum of two CT scans, and a minimum of 5 blood pressure and heart rate recordings in the first 24 hours following TEVAR were included in this sample, resulting in 31 subjects. Data used for this analysis included demographics, all available blood pressure data, and all available heart rate data. The blood pressure and heart rate data were cleaned (removed outliers/inaccurate values), and pulse pressure (PP) was calculated as the difference between SBP and diastolic blood pressure (DBP). Preoperative aortic geometry measures were calculated using preoperative CT scans.^5^ Two patient subgroups are derived from the main cohort to analyze hemodynamic predictors of outcomes following TEVAR: a failed TEVAR cohort of 14 patients and a successful TEVAR cohort of 17 patients. Outcomes were determined by whether a secondary surgical intervention is required more than 30 days after the initial TEVAR procedure. Since this data cohort is retrospective, all reintervention decisions were made by the primary surgeon. The most frequent reasons for reintervention following TEVAR were ongoing false lumen (FL) expansion and type 1A leak.

### Statistical Analysis

Statistical analyses were performed using Python 3.9, utilizing the packages numpy, pandas, SciPy, statsmodels, and scikit-learn. Descriptive statistics (mean, standard deviation, range, and kurtosis) were calculated for all study variables, including SBP, DBP, PP, and HR. Pearson correlations and two-tailed T-tests were calculated between basic statistics and TEVAR outcomes. Significance for this exploratory analysis was set at p = .10.

Next, the time-series data of SBP, DBP, and PP was subjected to the three stationarity tests stationarity tests Augmented Dickey-Fuller test (ADF), Kwiatkowski-Phillips-Schmidt-Shin (KPSS) and Auto-correlation function (ACF) in the Python package statsmodels 0.14.0 to investigate the stationarity of the data. The ADF tests for the presence of serial (autoregressive) correlation in time series data *y_t_* by performing a linear regression of Δ*y_t_* (*y_t_* - *y_t_*_−1_) against t, *y_t_*_−*p*_ where *y_t_*_−*p*_ is a p-order shifted version of *y_t_* such that:

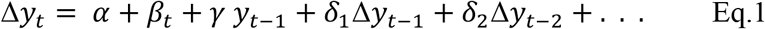

The null hypothesis is that the data are non-stationary and the coefficients of the equation (Eq.1) are all equal to zero; if not, then we have a stationary process.^15, 16^ KPSS also tests for stationarity^17–21^, but it assumes the presence of a trend in the data and tests for the stationarity around that trend. ACF measures the linear relationship between data points at time t and previous times.^22–24^

### Machine Learning Model

A machine learning model was developed to predict the outcome of TEVAR surgery based on features engineered from the CT scan, blood pressure, and HR data including descriptive statistics and stationarity-derived features. The following descriptive statistics: mean, standard deviation, kurtosis, and range were calculated for the time series data of SBP, DBP, PP, and HR for each patient. Similarly, the p-values of three stationarity tests (ADF, KPSS, and ACF) were calculated for each time series data for each patient. The p-value features derived from those tests were further engineered to binary features that indicate whether the outcome of the test was statistically significant or not at a cutoff of α = 0.01.

Using scikit-learn, logistic regression models were performed to predict a binary target outcome indicating whether the patient had a favorable TEVAR outcome or not. All the features were standardized features by removing the mean and scaling to unit variance except for binary features. The model using only the two CT scan-derived features (Mean Curvedness (MC) and Total Curvature Deviation (*δK*)) was used as a baseline model test for the added discriminative ability of the blood pressure-derived features, and five-fold cross-validation was used to validate the model results.

## Results

A total of 560 blood pressures across 31 patients were available for analysis with an average of 18.1 ± 6.0 readings available for each subject (range 5 – 29). For HR, the total number of readings was 680 with an average of 21.9 ± 7.8 per subject (range 7 – 38).

### Correlational Analysis

Patients with successful post-operative outcomes tended to have a higher mean PP after surgery with a mild positive Pearson correlation of 0.29. Moreover, patients with successful outcomes tend to have higher standard deviations and range values for their DBP time series with mild positive Pearson correlations of 0.33 and 0.31 respectively (Figure 1). The standard deviation (STD) and range of DBP were both significant with p values of 0.07 and 0.08 respectively while all other the t-tests came as non-significant indicating that basic statistics are not sufficient to discriminate between TEVAR patients of successful vs unsuccessful outcomes. For HR data, only the standard deviation showed significant correlation with the outcome with a p value of 0.07 (α = 0.10) and a weak negative correlation of -0.33.

**Figure 1:**
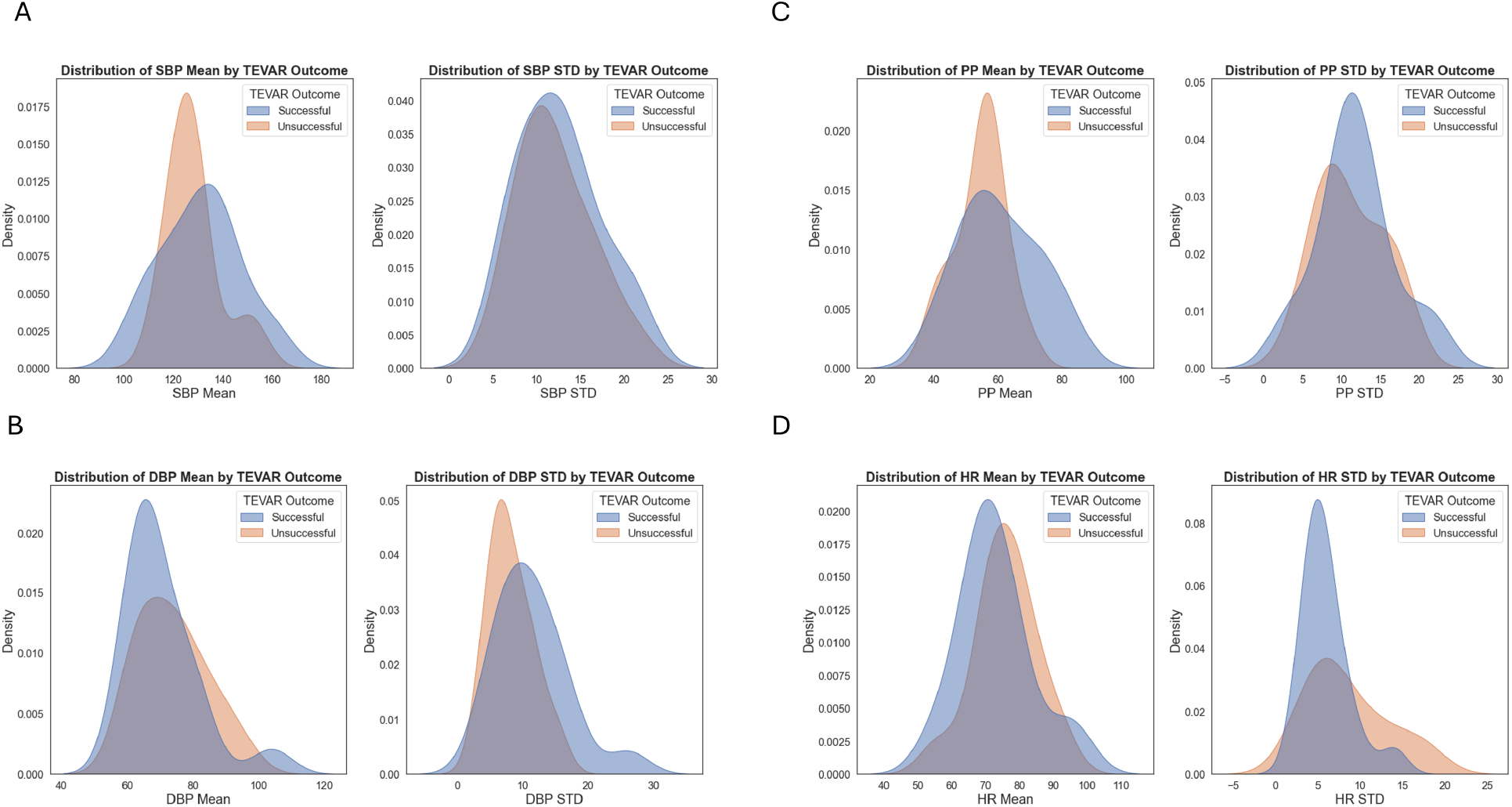
Kernel Density Estimation (KDE) plots illustrating the distributions of mean and standard deviation for A) systolic blood pressure (SBP), B) diastolic blood pressure (DBP), C) pulse pressure (PP), D) heart rate in patients with successful versus unsuccessful outcomes from the 24h following TEVAR surgery. The shaded areas represent the density of measurements for each group, highlighting distributions of flood pressure metrics in the two patient populations.

### Stationarity Tests

Stationarity in time series analysis refers to a property where the statistical characteristics of a time series, such as its mean, variance, and autocorrelation, remain constant over time (Figure 2). A stationary time series does not exhibit trends, seasonal effects, or other patterns that change over time. The two most common stationarity tests are KPSS and ADF while the ACF test helps to identify patterns, such as trends or seasonality, by showing how data points are related to past values.

**Figure 2:**
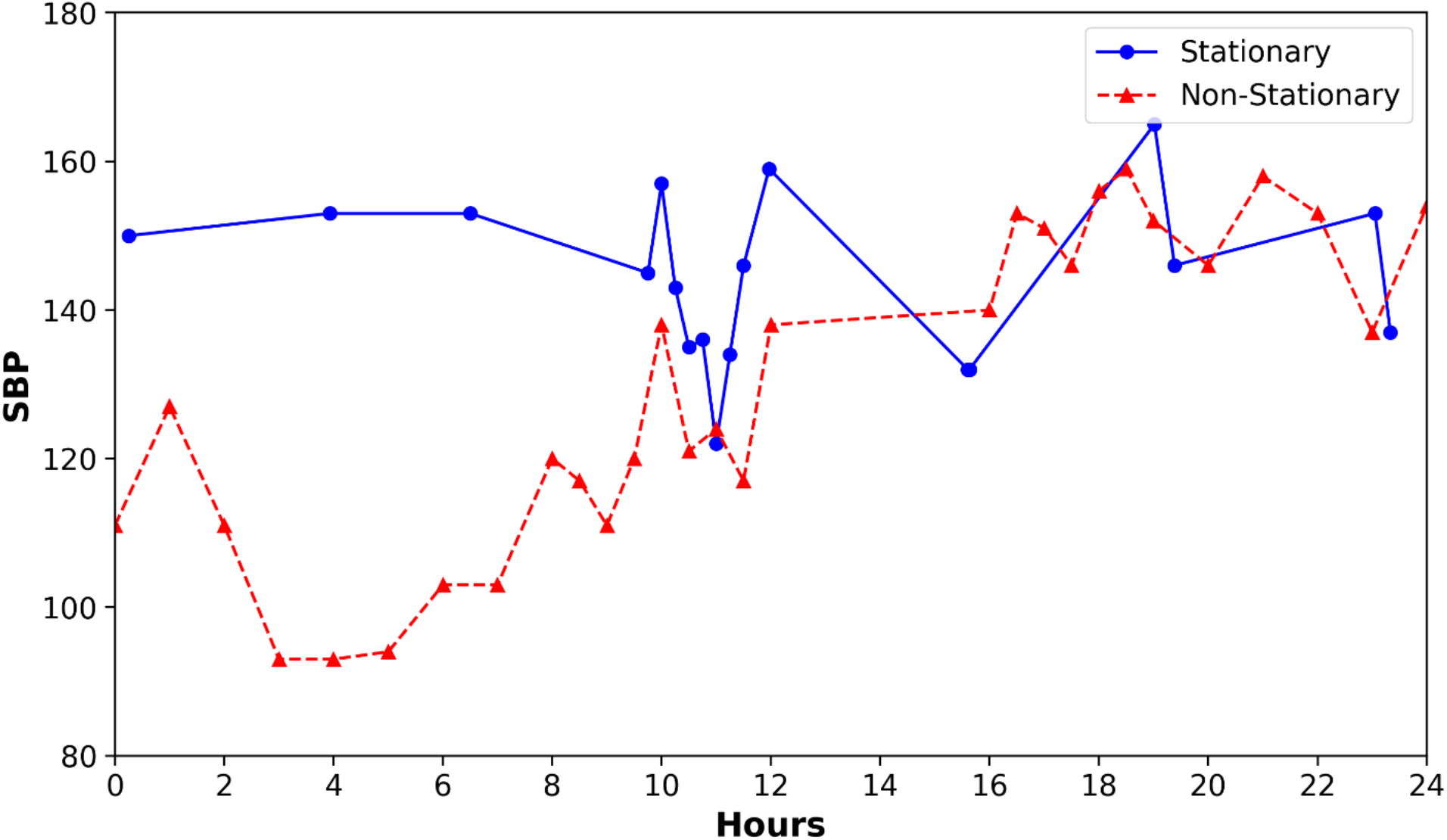
Representative Examples of Systolic Blood Pressure (SBP) Timeseries Data Over 24 hours Post-TEVAR: The blue solid line illustrates the SBP of a successful TEVAR patient showing stationary behavior, while the red dotted line depicts the SBP of an unsuccessful TEVAR patient with non-stationary behavior.

The p values of the three stationarity tests (ADF, ACF, KPSS) were calculated for SBP, DPB, PP, and HR time-series data of each patient, and Pearson correlations were calculated between those p values and the outcome variable. It was found that the p value of the KPSS test for SBP had a low positive correlation with the output variable of 0.3 while the p value of the ADF test for SBP had a low negative correlation with the ADF test equal to -0.31. Moreover, when a two-tailed t-test was applied to those p values between the two patient groups. It was found that the p values of the ADF test on SBP values are significantly lower in the patients with successful outcomes (p-value = 0.0844) at a significance level of (α = 0.1). For HR, stationarity was found to be uncorrelated with the outcome.

To further investigate possible correlations between stationarity of HR and BP data and the outcome, the calculated p-values were further engineered to binary features that indicate whether the outcome of the test was statistically significant or not at a cutoff of (α = 0.05). Followingly, the Fisher’s Exact test^25^ odds ratio and p-values were calculated between those new binary features and the outcome variable. The Fisher’s Exact test was performed instead of the chi-square test due to the binary nature of the engineered significance variables, and the presence of call frequencies less than 5 in some of the categories. The odds ratio of the Fisher’s Exact test showed the correlation between the significance of the ADF test on SBP time-series data and the outcome variable with an odds ratio of 2.2 which implies a positive association. Also, the odds ratio between the significance of the ACF test on PP and the outcome variable was 3.3 indicating a positive correlation. Similarly, Fisher’s Exact test between the significance of the ADF test on PP time-series data and the outcome variable showed an odds ratio of 0.24 suggesting a relatively strong negative association, however, the p-value did not show any significance (p-value = 0.197). The significance of the ADF test on SBP was also investigated at a stricter significance level of (α = 0.01), the binary features produced at this cutoff had an odds ratio of 5.3 with the output variable and a p-value of 0.0680 using Fisher’s exact test.

### SBP Stationarity Tests

The previous results indicate that the stationarity of the SBP time-series data can be a useful predictor of the TEVAR outcome given that the p values of the ADF test on SBP and the p values of the KPSS test on SBP showed a correlation with the output variable. Based on this observation, the patients were categorized into four categories based on the significance of the KPSS and the ADF test on SBP:

- **Strictly stationary:** ADF and KPSS both suggest stationarity.
- **Trend-stationary:** ADF suggests stationarity, while KPSS suggests non-stationarity.
- **Difference-stationary:** ADF suggests non-stationarity, while KPSS suggests stationarity.
- **Non-stationarity**: ADF and KPSS both suggest non-stationarity.

Fisher’s Exact odds ratios suggest that the odds of having a successful TEVAR outcome with a strictly stationary SBP are 3.3 times higher than the odds of having an unsuccessful TEVAR outcome. Also, the percentage of patients with strictly stationary SBP in the favorable TEVAR outcome group was almost twice the percentage in the unsuccessful TEVAR outcome group (Figure 3B). Those results indicate that combining the significance of two of the stationarity tests (KPSS and ADF) on SBP can be a good predictor of the TEVAR outcome. Cross counts of those two tests are shown in Figure 3A.

**Figure 3:**
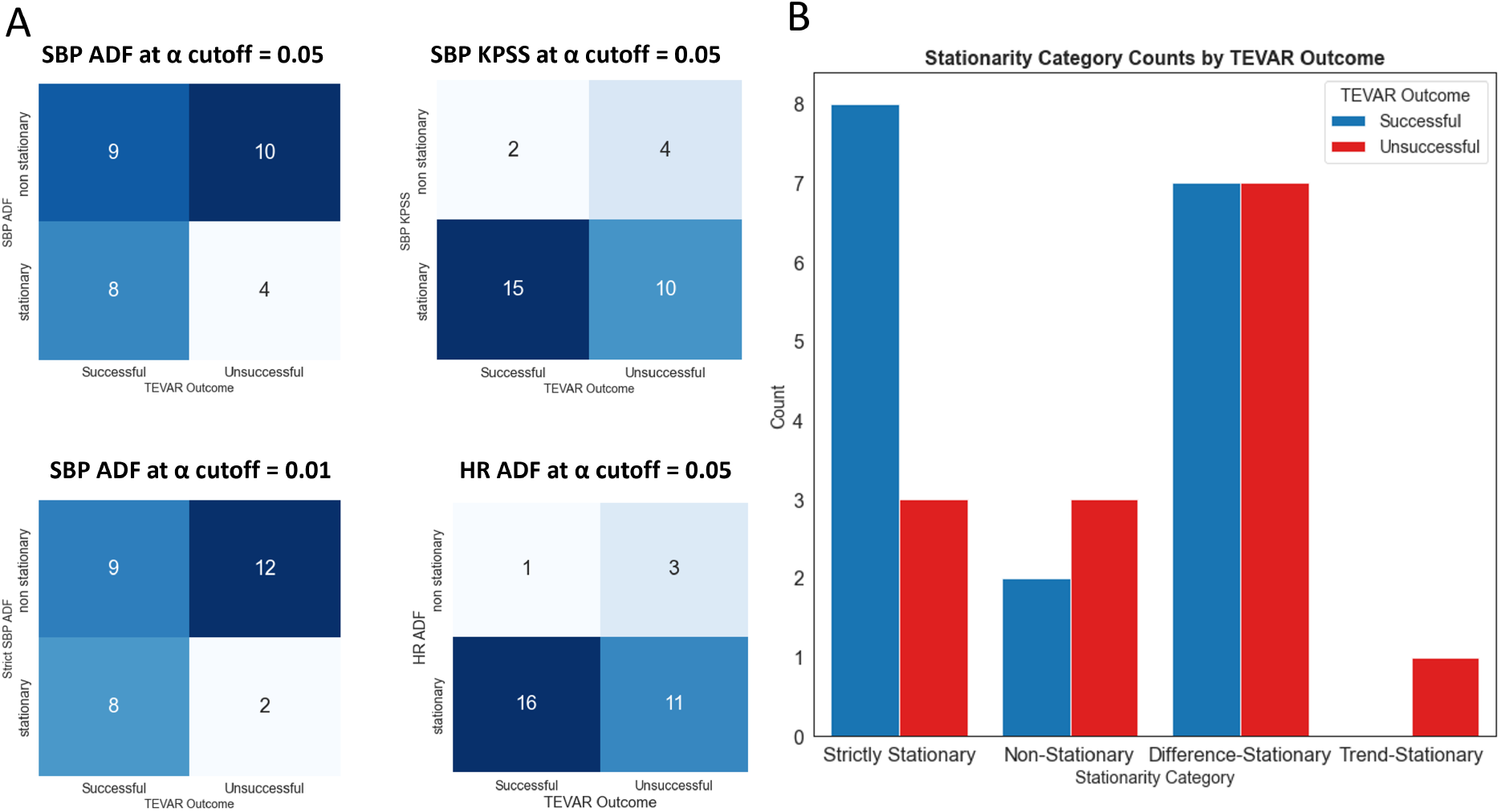
A) Cross counts of stationarity based on the Adfuller and KPSS tests for SBP and HR. B) Percentage of patients of each TEVAR outcome group that fall into SBP stationarity categories.

### Machine Learning Models

The best features were used to predict the outcome variable in a logistic regression model to calculate the regression coefficients and select the best set of features. At first glance, it was clear that the binary significance feature of the ADF test on SBP was the best blood pressure-derived feature to boast the practicability of the CT scan-derived features (Figure 4). Furthermore, two other BP features (the standard deviation of the DPB, the p-value of the KPSS on SBP) and the std of HR boosted the model performance when combined with CT scan-derived features and the significance feature of the ADF test on SBP leading to a model with six features that achieved the best five-fold cross-validation accuracy of 96.67% in predicting TEVAR outcome. Full results are shown in Table 1.

**Figure 4:**
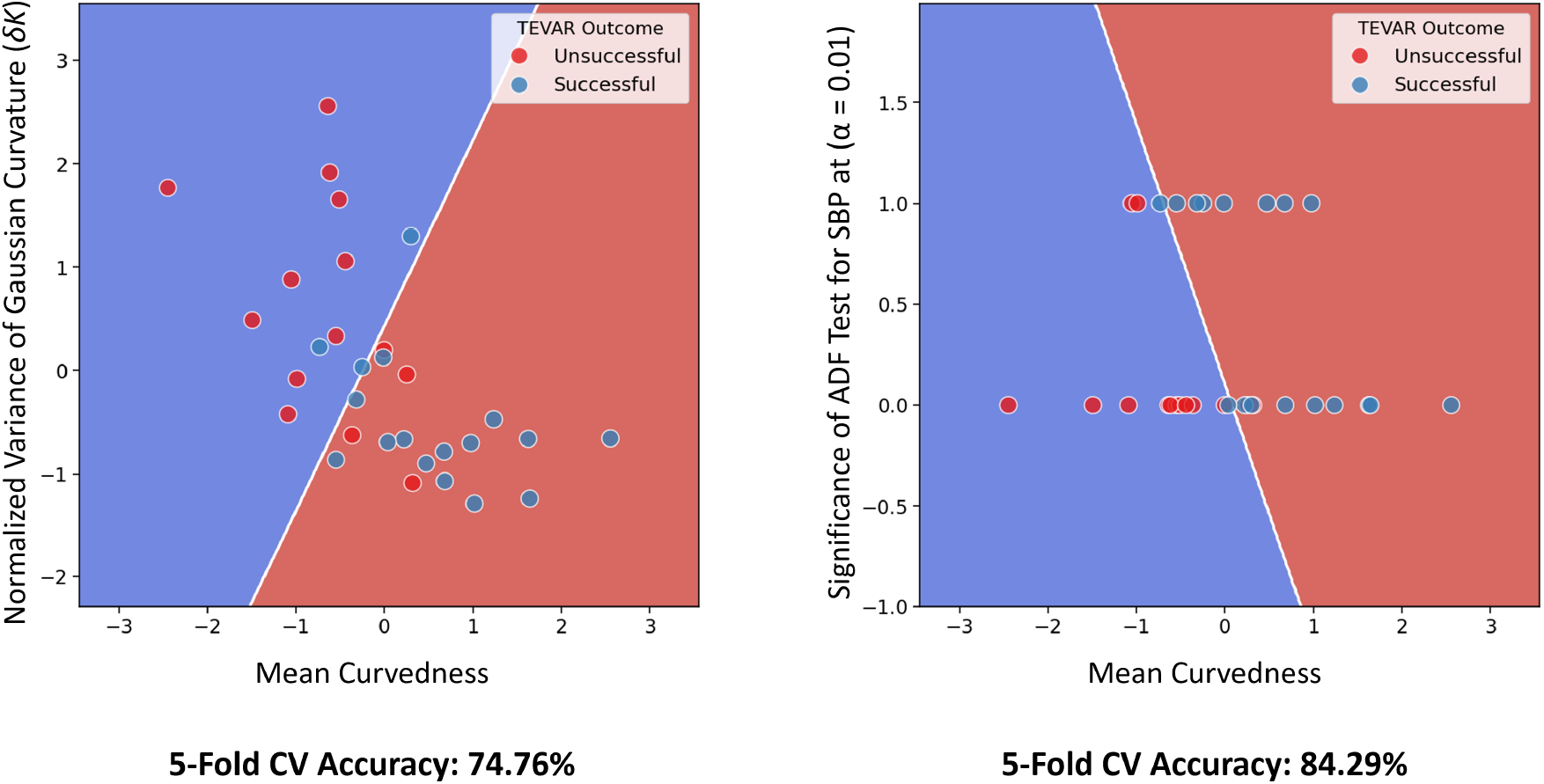
Decision Boundaries Comparison for Logistic Regression Models Classifying Successful and Unsuccessful TEVAR Patients: *Left*: Decision boundary of the logistic regression model trained using aortic CTA imaging features: Mean Curvedness (MC) and Normalized Variance of Gaussian Curvature (δK). *Right*: Decision boundary of the logistic regression model trained using an aortic CTA imaging feature (Mean Curvedness) combined with a post-TEVAR systolic blood pressure binary feature. This feature represents the Significance of SBP, determined by the Augmented Dickey-Fuller (ADF) stationarity test at a significance level of α = 0.01 (where 1 indicates stationary SBP and 0 indicates non-stationary SBP).

**Table 1:** Results of logistic regression models predicting TEVAR outcomes based on different features derived from CT scans, BP, and HR data in the 24 hours post-surgery. (Abbreviations: MC: Mean Curvedness, *δK*: Normalized Variance of Gaussian Curvature, SAS: strict significance of the ADF test of SBP at α = 0.01, SBP KPSS: p-values of the KPSS test on SBP data).

| *Features Used* | **5-fold CV accuracy** |
| --- | --- |
| MC + $\delta K$ (*CT only baseline*) | 74.76% |
| MC + $\delta K$ + SAS | 83.81% |
| MC + $\delta K$ + SAS + DBP std | 90.95% |
| MC + $\delta K$ + SAS + DBP std + SBP KPSS | 93.33% |
| MC + $\delta K$ + SAS + DBP std + SBP KPSS +HR std | **96.67%** |
| SAS + DBP std + SBP KPSS (*BP metrics*) | 73.33% |

## Discussion

Hemodynamic factors, along with geometric, morphological, biomechanical, and genetic factors, underlie the development and progression of aortic dissection. While hemodynamics are being investigated as biomarkers for prognosis and treatment decisions, the majority of those studies focus on hemodynamics within the aorta itself and not clinical data that is readily available.^2, 3, 26–28^ Furthermore, the complex interplay between systemic hemodynamic factors and aortic dissection geometry and morphology is incompletely understood. In this study, we found that adding hemodynamic metrics creates a novel multi-dimensional aortic geometric-physiologic feature space and improves our ability to predict post-TEVAR outcomes.

The variable that best contributed to this model was the SBP ADF stationarity measure, with nonstationarity being associated with worse outcomes. A common cause of nonstationarity is the presence of a trend, which means the mean of the series changes over time, affecting its variability. Long-term systolic variability has previously been identified as a risk factor for cardiovascular events such as stroke and cardiovascular mortality.^9, 12^ While these studies have historically used visit-to-visit data, recent work in other vascular diseases have utilized EMR data to look at the impact of systolic variability. In fact, increased systolic variability during hospitalization has been associated with worse clinical outcomes in stroke^29^, increased risk of mortality among patients with peripheral artery disease^30^ and renal function decline and cardiovascular events in coronary artery disease.^31^ These findings, along with the findings of the present study, indicate that temporal trends in SBP must be evaluated to better identify patients at risk of poor outcomes.

Diastolic blood pressure standard deviation also contributed to improvements in model performance. Standard deviations of hemodynamic readings have frequently been used to characterize variability, demonstrating variations from a patient’s baseline and not temporal trends like stationarity. Variability in DBP, along with SBP, may result in inappropriate remodeling of the aorta due to fatigue.^32^ The fatigue of arteries is a chronic failure process caused by numerous cyclic blood pressures. If the amount of pressure during both systole and diastole is inconsistent, the rate of arterial crack growth accelerates.^33^ This association between cyclical loading and vessel fatigue may underpin the connection between blood pressure variability and the adverse outcomes in aortic dissection demonstrated in this study.

Similarly, the standard deviation of the heart rate over this 24 hour period improved the model’s ability to predict TEVAR outcomes. Heart rate variability metrics reflect regulation of multiple autonomic processes, including vascular tone and blood pressure.^34^ A healthy heart exhibits variability at baseline, providing the flexibility to cope with changing biologic demands and environments. However, changes in that expected complexity can indicate a pathological condition.^35^ For example, heart rate variability has been identified as a key predictor for adverse cardiovascular outcomes including in-hospital cardiac arrest.^12, 36, 37^ There are multiple ways to capture heart rate variability, including standard deviations of beat-to-beat interval changes;^38^ while our measure is atypical as it uses the data available in the medical record, the signal indicates that heart rate variability should be more closely examined in AD.

These findings emphasize the importance of appropriately collecting and using temporal measurements. Postoperatively, we generally have frequent data readings on hemodynamic parameters; yet, the majority of care decisions are made based on singular values or trends that can be evaluated by the naked eye. In order to fully harness the data we collect, we must convert raw, high-frequency data into intuitive visual formats that highlight clinical trajectories over time. There are efforts underway to maximize the use of this data, including the MIMIC II relational database that captures continuous data from ICU patient monitors.^39^ Similarly, real-time clinical support tools using AI have been proposed to assess for patterns that may predict adverse clinical outcomes.^40^ However, more work must be done to build trust in healthcare AI models to ensure clinican adoption and thereby best use this invaluable data.^41^

Several factors may limit the generalizability of these findings. First, this is a small cohort from a single center. Additionally, this study used retrospective data; despite careful data cleaning to remove values that are hemodynamically impossible, there is the potential that errors existed within the dataset. This study also did not account for medical management, including medications such as vasopressors and antihypertensives that could impact both blood pressure and heart rate. Future research should take these factors into consideration and evaluate the impact of medical management on hemodynamic variability on AD and TEVAR outcomes. Finally, this study utilized an atypical measure of heart rate variability by looking at the standard deviation of all heart rate measures captured in the electronic health record. Gold standard measures of heart rate variability, including beat-to-beat measures, should be considered in a future prospective cohort to gain additional insight and compare to the novel approach we used in this study.

## Conclusion

Post-TEVAR behaviors of hemodynamic variables are critical predictors of TEVAR success, with higher DBP standard deviation and strict stationarity of SBP significantly associated with favorable outcomes. Combining these time-series metrics with preoperative aortic geometry and HR standard deviation in predictive models enhanced accuracy to 96.67%, underscoring the value of integrating physiological and imaging-derived features. These findings highlight the importance of comprehensive blood pressure and heart rate monitoring in the critical 24-hour post-TEVAR period to improve patient management and outcomes.

## Data Availability

All data produced in the present study are available upon reasonable request to the authors.

## Funding

We acknowledge the support of the National Institutes of Health, NHLBI, R01HL159205 to LP. The Center for Research Informatics is funded by the Biological Science Division at The University of Chicago with additional funding provided by the Institute for Translational Medicine, CTSA grant number ULITR000430 from NIH. The funders had no role in study design, data collection and analysis, decision to publish, or preparation of the manuscript.

## Notes

### Competing Interest Statement

The authors have declared no competing interest.

### Author Declarations

The IRB of the University of Chicago gave ethical approval for this work.

## References

1. Mussa FF, Horton JD, Moridzadeh R, Nicholson J, Trimarchi S, Eagle KA. Acute Aortic Dissection and Intramural Hematoma: A Systematic Review. JAMA. 2016;316(7):754–63. doi: 10.1001/jama.2016.10026. PubMed PMID: 27533160.

2. Marlevi D, Sotelo JA, Grogan-Kaylor R, Ahmed Y, Uribe S, Patel HJ, Edelman ER, Nordsletten DA, Burris NS. False lumen pressure estimation in type B aortic dissection using 4D flow cardiovascular magnetic resonance: comparisons with aortic growth. J Cardiovasc Magn Reson. 2021;23(1):51. Epub 20210513. doi: 10.1186/s12968-021-00741-4. PubMed PMID: 33980249; PMCID: PMC8117268.

3. Cheng Z, Tan FP, Riga CV, Bicknell CD, Hamady MS, Gibbs RG, Wood NB, Xu XY. Analysis of flow patterns in a patient-specific aortic dissection model. J Biomech Eng. 2010;132(5):051007. doi: 10.1115/1.4000964. PubMed PMID: 20459208.

4. Chi Q, He Y, Luan Y, Qin K, Mu L. Numerical analysis of wall shear stress in ascending aorta before tearing in type A aortic dissection. Comput Biol Med. 2017;89:236–47. Epub 20170801. doi: 10.1016/j.compbiomed.2017.07.029. PubMed PMID: 28843154.

5. Liu M, Dong H, Mazlout A, Wu Y, Kalyanasundaram A, Oshinski JN, Sun W, Elefteriades JA, Leshnower BG, Gleason RL, Jr. The role of anatomic shape features in the prognosis of uncomplicated type B aortic dissection initially treated with optimal medical therapy. Comput Biol Med. 2024;170:108041. Epub 20240129. doi: 10.1016/j.compbiomed.2024.108041. PubMed PMID: 38330820; PMCID: PMC10932856.

6. Isselbacher EM, Preventza O, Hamilton Black J, 3rd, Augoustides JG, Beck AW, Bolen MA, Braverman AC, Bray BE, Brown-Zimmerman MM, Chen EP, Collins TJ, DeAnda A, Jr., Fanola CL, Girardi LN, Hicks CW, Hui DS, Schuyler Jones W, Kalahasti V, Kim KM, Milewicz DM, Oderich GS, Ogbechie L, Promes SB, Gyang Ross E, Schermerhorn ML, Singleton Times S, Tseng EE, Wang GJ, Woo YJ, Peer Review Committee M. 2022 ACC/AHA Guideline for the Diagnosis and Management of Aortic Disease: A Report of the American Heart Association/American College of Cardiology Joint Committee on Clinical Practice Guidelines. Circulation. 2022;146(24):e334–e482. Epub 20221102. doi: 10.1161/CIR.0000000000001106. PubMed PMID: 36322642; PMCID: PMC9876736.

7. Hutcheon JA, Chiolero A, Hanley JA. Random measurement error and regression dilution bias. BMJ. 2010;340:c2289. Epub 20100623. doi: 10.1136/bmj.c2289. PubMed PMID: 20573762.

8. Wagner CD, Nafz B, Persson PB. Chaos in blood pressure control. Cardiovasc Res. 1996;31(3):380–7. PubMed PMID: 8681325.

9. Dasa O, Smith SM, Howard G, Cooper-DeHoff RM, Gong Y, Handberg E, Pepine CJ. Association of 1-Year Blood Pressure Variability With Long-term Mortality Among Adults With Coronary Artery Disease: A Post Hoc Analysis of a Randomized Clinical Trial. JAMA Netw Open. 2021;4(4):e218418. Epub 20210401. doi: 10.1001/jamanetworkopen.2021.8418. PubMed PMID: 33914047; PMCID: PMC8085725.

10. Anisetti B, Youssef H, Elkhair AM, Lin MP. Association between within-visit blood pressure variability, stroke, coronary heart disease, and cardiovascular mortality. Neurol Neurochir Pol. 2023;57(5):423–9. Epub 20230901. doi: 10.5603/pjnns.94839. PubMed PMID: 37655640.

11. Benetos A, Thomas F, Bean K, Albaladejo P, Palatini P, Guize L. Resting heart rate in older people: a predictor of survival to age 85. J Am Geriatr Soc. 2003;51(2):284–5. doi: 10.1046/j.1532-5415.2003.51080.x. PubMed PMID: 12558738.

12. Zeng R, Wang Z, Cheng W, Yang K. Visit-to-Visit Heart Rate Variability Is Positively Associated With the Risk of Adverse Cardiovascular Outcomes. Front Cardiovasc Med. 2022;9:850223. Epub 20220307. doi: 10.3389/fcvm.2022.850223. PubMed PMID: 35321108; PMCID: PMC8936423.

13. Hartl J, Sander D, Ikenberg ID, Schmidt G, Hemmer B, Wunderlich S, Nimmrichter B, Purner K, Hapfelmeier A. Is the association between visit-to-visit heart rate variability and cardiovascular disease mediated by arteriopathy as measured by carotid intima-media thickness? J Stroke Cerebrovasc Dis. 2025;34(8):108348. Epub 20250514. doi: 10.1016/j.jstrokecerebrovasdis.2025.108348. PubMed PMID: 40379229.

14. Khabaz K, Yuan K, Pugar J, Jiang D, Sankary S, Dhara S, Kim J, Kang J, Nguyen N, Cao K, Washburn N, Bohr N, Lee CJ, Kindlmann G, Milner R, Pocivavsek L. The geometric evolution of aortic dissections: Predicting surgical success using fluctuations in integrated Gaussian curvature. PLoS Comput Biol. 2024;20(2):e1011815. Epub 20240202. doi: 10.1371/journal.pcbi.1011815. PubMed PMID: 38306397; PMCID: PMC10866512.

15. Cheung YW, Lai KS. Lag Order and Critical Values of a Modified Dickey-Fuller Test. Oxford Bulletin of Economics and Statistics. 1995;57(3):411–9.

16. Dickey DA, Fuller WA. Distribution of the estimators for autoregressive time series with a unit root. Journal of the American statistical association. 1979;74(366a):427–31.

17. Schwert GW. Tests for unit roots: A Monte Carlo investigation. Journal of business & economic statistics. 2002;20(1):5–17.

18. Hobijn B, Franses PH, Ooms M. Generalizations of the KPSS-test for stationarity. Statistica Neerlandica. 2004;58(4):483–502.

19. Newey WK, West KD. Automatic lag selection in covariance matrix estimation. The review of economic studies. 1994;61(4):631–53.

20. Andrews DWK. Heteroskedasticity and autocorrelation consistent covariance matrix estimation. Econometrica: Journal of the Econometric Society. 1991:817–58.

21. Kwiatkowski D, Phillips PCB, Schmidt P, Shin Y. Testing the null hypothesis of stationarity against the alternative of a unit root: How sure are we that economic time series have a unit root? Journal of econometrics. 1992;54(1-3):159–78.

22. Brockwell PJ, Davis RA. Nonstationary and seasonal time series models. Introduction to time series and forecasting: Springer; 2016. p. 157–93.

23. Brockwell PJ, Davis RA. Time series: theory and methods: Springer science & business media; 2009.

24. Parzen E. On spectral analysis with missing observations and amplitude modulation. Sankhyā: The Indian Journal of Statistics, Series A. 1963:383–92.

25. Fisher RA. Mathematics of a lady tasting tea. The world of mathematics. 1956;3(part 8):1514–21.

26. Burris NS, Nordsletten DA, Sotelo JA, Grogan-Kaylor R, Houben IB, Figueroa CA, Uribe S, Patel HJ. False lumen ejection fraction predicts growth in type B aortic dissection: preliminary results. Eur J Cardiothorac Surg. 2020;57(5):896–903. doi: 10.1093/ejcts/ezz343. PubMed PMID: 31821480; PMCID: PMC8453376.

27. Xu H, Xiong J, Han X, Mei Y, Shi Y, Wang D, Zhang M, Chen D. Computed tomography-based hemodynamic index for aortic dissection. J Thorac Cardiovasc Surg. 2021;162(2):e165–e76. Epub 20200220. doi: 10.1016/j.jtcvs.2020.02.034. PubMed PMID: 32217023.

28. Zimmermann J, Baumler K, Loecher M, Cork TE, Marsden AL, Ennis DB, Fleischmann D. Hemodynamic effects of entry and exit tear size in aortic dissection evaluated with in vitro magnetic resonance imaging and fluid-structure interaction simulation. Sci Rep. 2023;13(1):22557. Epub 20231218. doi: 10.1038/s41598-023-49942-0. PubMed PMID: 38110526; PMCID: PMC10728172.

29. Guo ZN, Qu Y, Reziya H, Liu J, Yan XL, Zhang P, Zhang PD, Qi S, Yang Y. Which Parameters of Beat-to-Beat Blood Pressure Best Predict Poor In-Hospital Outcome in Spontaneous Intracerebral Hemorrhage? Front Aging Neurosci. 2020;12:603340. Epub 20201119. doi: 10.3389/fnagi.2020.603340. PubMed PMID: 33328978; PMCID: PMC7710867.

30. Suzuki M, Saito Y, Kitahara H, Saito K, Takahara M, Himi T, Kobayashi Y. Impact of in-hospital blood pressure variability on clinical outcomes in patients with symptomatic peripheral arterial disease. Hypertens Res. 2021;44(8):1002–8. Epub 20210413. doi: 10.1038/s41440-021-00648-8. PubMed PMID: 33850306.

31. Saito K, Saito Y, Kitahara H, Nakayama T, Fujimoto Y, Kobayashi Y. In-Hospital Blood Pressure Variability: A Novel Prognostic Marker of Renal Function Decline and Cardiovascular Events in Patients with Coronary Artery Disease. Kidney Blood Press Res. 2020;45(5):748–57. Epub 20201007. doi: 10.1159/000509291. PubMed PMID: 33027787.

32. Chaudhry HR, Bukiet B, Davis A, Ritter AB, Findley T. Residual stresses in oscillating thoracic arteries reduce circumferential stresses and stress gradients. J Biomech. 1997;30(1):57–62. doi: 10.1016/s0021-9290(97)81292-4.

33. Versluis A, Bank AJ, Douglas WH. Fatigue and plaque rupture in myocardial infarction. J Biomech. 2006;39(2):339–47. Epub 20050118. doi: 10.1016/j.jbiomech.2004.10.041. PubMed PMID: 16321636.

34. Rajendra Acharya U, Paul Joseph K, Kannathal N, Lim CM, Suri JS. Heart rate variability: a review. Med Biol Eng Comput. 2006;44(12):1031–51. Epub 20061117. doi: 10.1007/s11517-006-0119-0. PubMed PMID: 17111118.

35. ChuDuc H, NguyenPhan K, NguyenViet D. A Review of Heart Rate Variability and its Applications. APCBEE Procedia. 2013;7:80–5. doi: 10.1016/j.apcbee.2013.08.016.

36. Lee H, Yang HL, Ryu HG, Jung CW, Cho YJ, Yoon SB, Yoon HK, Lee HC. Real-time machine learning model to predict in-hospital cardiac arrest using heart rate variability in ICU. NPJ Digit Med. 2023;6(1):215. Epub 20231123. doi: 10.1038/s41746-023-00960-2. PubMed PMID: 37993540; PMCID: PMC10665411.

37. Brinza C, Floria M, Scripcariu DV, Covic AM, Covic A, Popa IV, Statescu C, Burlacu A. Heart Rate Variability in Acute Myocardial Infarction: Results of the HeaRt-V-AMI Single-Center Cohort Study. J Cardiovasc Dev Dis. 2024;11(8). Epub 20240822. doi: 10.3390/jcdd11080254. PubMed PMID: 39195162; PMCID: PMC11355001.

38. Shaffer F, Ginsberg JP. An Overview of Heart Rate Variability Metrics and Norms. Front Public Health. 2017;5:258. Epub 20170928. doi: 10.3389/fpubh.2017.00258. PubMed PMID: 29034226; PMCID: PMC5624990.

39. Saeed M, Lieu C, Raber G, Mark RG, editors. MIMIC II: a massive temporal ICU patient database to support research in intelligent patient monitoring. Computers in cardiology; 2002: IEEE.

40. Cannesson M, Hofer I, Rinehart J, Lee C, Subramaniam K, Baldi P, Dubrawski A, Pinsky MR. Machine learning of physiological waveforms and electronic health record data to predict, diagnose and treat haemodynamic instability in surgical patients: protocol for a retrospective study. BMJ open. 2019;9(12):e031988.

41. Arleo A, Chen AT, Gotz D, Kandaswamy S, Bernard J. Reflections on interactive visualization of electronic health records: past, present, future. J Am Med Inform Assoc. 2024;31(11):2423–8. doi: 10.1093/jamia/ocae249. PubMed PMID: 39429003; PMCID: PMC11491741.

